# Do Calibrated-Probability Claims Hold Up in Clinical Diagnosis? A Pre-registered Evaluation of a Decision Model Against Frontier Language Models

**DOI:** 10.64898/2026.10.09.26365153

**Authors:** Avnish Deobhakta, Jonathan Chang, Royce W.S. Chen, Sujai Jaipalli, Alexander Pinhas, Tak Yee Tania Tai, Richard B. Rosen

## Abstract

**Background:** A new class of AI “decision models” returns a probability for every option in a single pass and is marketed as calibrated, which invites use in clinical triage. Whether those probabilities hold up on clinical diagnosis has not been independently tested.

**Objective:** To test the diagnostic accuracy, calibration, and probabilistic coherence of a decision model (Jev 1.13, TypeSafe AI) against frontier language models, with and without reasoning.

**Methods:** In a pre-registered study (OSF, registered 26 September 2026), we compared Jev with Claude Opus 5.5 (adaptive reasoning, always on) and Claude Opus 5 (reasoning disabled) on three case sets: the 45 Semigran symptom-checker vignettes, 1,000 stratified synthetic patients from DDXPlus with reference differentials, and 80 simulated ophthalmic complaints (40 diagnoses at two levels of detail). Each model returned a probability for every diagnosis on a fixed list. Outcomes were top-1 and top-3 accuracy, expected calibration error (ECE), Brier score, the rate of exactly zero probability on the true diagnosis, sensitivity to option order, behavior on uninformative inputs, and behavior when the true diagnosis was absent.

**Results:** On DDXPlus, top-1 accuracy was 72.0% for Jev, 79.5% for Opus 5 without reasoning, and 87.4% for Opus 5.5 (both comparisons with Jev P<.001). Jev assigned exactly zero probability to the true diagnosis in 62 of 1,000 cases (6.2%); neither comparator did so in any case. Calibration error did not differ between Jev and the registered comparator (ECE difference 0.007, 95% CI -0.026 to 0.038). Offered an explicit “none of these” option, Jev declined to diagnose all 30 uninformative inputs; without that option, it assigned a mean 58% to a single diagnosis. Order sensitivity was small in all models. Given the option list with no patient information, Jev placed up to 96% on a single diagnosis, while the comparators spread probability almost evenly. On the ophthalmic complaints, accuracy did not differ (Jev 88.8%, Opus 5 91.2%, Opus 5.5 88.8%), and Jev was better calibrated than the registered comparator (ECE difference -0.081, 95% CI -0.124 to -0.018).

**Conclusions:** The decision model matched frontier models on short, distinctive ophthalmic complaints but fell well behind on multi-symptom synthetic patients, including behind a comparator without reasoning, and there it excluded the correct diagnosis outright in about 1 in 16 cases. Its probabilities signaled uncertainty only when the question offered a way to abstain. Calibration claims for clinical decision models should be tested on clinical tasks, with explicit abstention options and zero-probability rates reported.

## Introduction

Health systems increasingly use AI to help triage patient portal messages and telephone calls as demand for physician access outgrows clinical capacity [1,2]. Probability outputs are what make a model usable for this purpose: software can act automatically when the model is confident and route the case to a clinician when it is not. Such a system works only if the probabilities are calibrated, meaning that a stated probability matches how often the model is actually right [3]. A model that assigns 90% to a diagnosis should be correct about 90% of the time, and a model given too little information to decide should be able to say so.

Large language models (LLMs) can be asked for probabilities, but they generate them as text, often after reasoning, at a cost of seconds and cents per answer. A new class of “decision models” takes a different route. Jev 1.13 (TypeSafe AI, released September 2026) generates no text: given a case and a list of options, it returns a probability for every option in one pass, at a small fraction of an LLM’s cost and latency. Its developer markets these probabilities as calibrated, the product of a training method aimed at calibration rather than fluent text [4]. On standardized clinical vignettes, general-purpose language models already diagnose and triage at levels approaching those of physicians [5,6], so a cheaper single-pass alternative would need to match that accuracy and calibration to be useful.

In the two weeks after release, independent developers tested Jev on customer-support routing, phishing detection, and licensing-exam questions, reporting nondeterminism, overconfidence on some question types, and above-chance accuracy from the answer options alone [7-9]. None of this work tested clinical diagnosis against a pre-specified plan, compared the model with frontier LLMs with reasoning both on and off, or examined how its probabilities fail. In medicine, how a model is wrong matters as much as how often.

This matters more in medicine than in the settings where such models are usually evaluated: a misrouted customer-support ticket costs time, but a missed diagnosis can cost vision or life.

We therefore ran a pre-registered evaluation of Jev against Claude Opus 5.5 and Claude Opus 5 on three diagnostic case sets. Our aim was not to rank products but to test whether calibrated-probability claims hold up on clinical diagnosis, and to identify the failure modes that matter for triage.

## Methods

### Design and registration

This was a pre-registered, cross-sectional evaluation of three AI models on published diagnostic case sets. The analysis plan, hypotheses, and violation thresholds were fixed before data collection and registered on the Open Science Framework (osf.io/fpmg3) [10] on 26 September 2026, before the first study model call. Two amendments made before any study data (scoring of log-loss, choice of Semigran input text) are included in the registration. Later deviations are reported at the end of the Results. No human participants were involved. Reporting follows the TRIPOD-LLM guideline for studies using large language models [11], and a completed checklist is provided as a supplementary file.

### Models

All models received the same case text, the same option keys in the same order, and the same instruction: “Which diagnosis best explains this patient’s presentation?” Each option was identified by a short text label derived from the diagnosis name (for example, acute_angle_closure_glaucoma) and described by the plain diagnosis name, with no definitions attached. Every response was stored with its timestamp, the provider’s resolved model version, and the code version that produced it.

**Table 1.** Models compared.

| Model | Version | Access | Reasoning | How probabilities were obtained |
| --- | --- | --- | --- | --- |
| Jev (TypeSafe AI) | jev-1.13- | OpenRouter | None (single pass) | Native: one Choice question per |
|  | 20260917 |  |  | case returns a probability for every option |
| Claude Opus 5.5 (Anthropic) | claude-opus-5-5 | Anthropic API | Adaptive, always on; cannot be disabled | JSON probability for every option key |
| Claude Opus 5 (Anthropic) | claude-opus-5 | Anthropic API | Disabled (effort high) | Same JSON elicitation as Opus 5.5 |

### Case sets

Semigran vignettes [12] were used in their published condensed form, which is the text the original study entered into symptom checkers and excludes test results that give away the answer. DDXPlus patients [13] were rendered from structured findings into prose with one fixed template. Zandi complaints [14] were used verbatim. The 30 uninformative inputs (for example, “I’ve been feeling a bit out of sorts lately. Not sure why.”) were written by a human collaborator and confirmed as untriageable by a clinician.

**Table 2.**
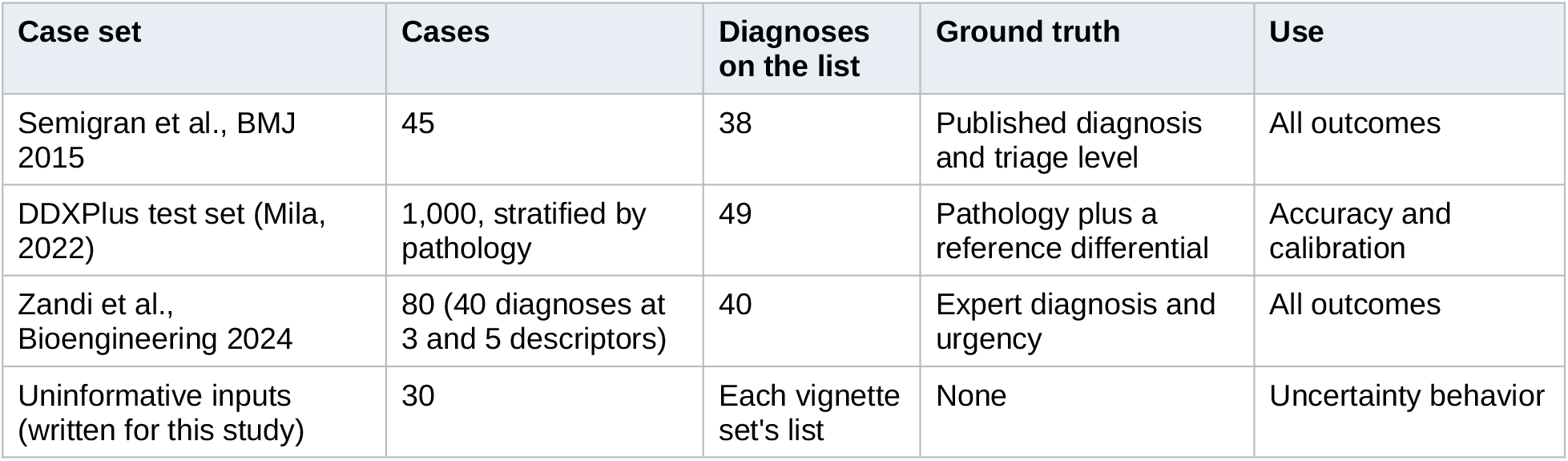
Case sets.

### Conditions

Each vignette was sent five times unchanged to measure run-to-run noise, five times with the options in random orders, and twice with its true diagnosis removed from the list (once with a “none of these” option added). DDXPlus cases were sent once each. Uninformative inputs were sent against each vignette set’s list, with and without “none of these.” An options-only baseline sent each list five times with no patient information. All random orders used a fixed seed.

### Outcomes

The registered primary outcomes were order invariance, independence from irrelevant options, and calibration. Accuracy was measured as top-1 and top-3. Calibration was measured by expected calibration error (10 equal-width bins on top-1 confidence) [15], multiclass Brier score, and log-loss after mixing each distribution with 1% uniform mass (so that exact zeros are scored identically for every model). Distance between two output distributions was measured by total variation distance (TVD). On DDXPlus, each model’s distribution was also compared with the reference differential by Jensen-Shannon divergence. We report the rate at which each model assigned exactly zero probability to the true diagnosis.

### Statistics

The case was the unit of analysis. Paired comparisons between models used exact McNemar tests for binary outcomes and case-level bootstrap intervals (10,000 resamples; 2,000 for calibration error) for continuous ones. Holm correction was applied across the registered primary comparisons. All code, data manifests, and results are version-controlled, and every result row records the commit that produced it.

## Results

Jev was less accurate than both comparators on the 1,000-case synthetic set and alone assigned zero probability to correct diagnoses there, but it matched them on the ophthalmic complaints and was the best calibrated model on that set.

### Accuracy and calibration

On DDXPlus, Jev’s top-1 accuracy was lower than that of Opus 5 without reasoning (only Jev correct in 45 cases, only Opus 5 correct in 120; P<.001) and Opus 5.5 (22 vs 176; P<.001). Opus 5.5 was also more accurate than Opus 5 (32 vs 111; P<.001). Jev’s Brier score was worse than both (difference vs Opus 5, 0.113; 95% CI 0.084 to 0.145). Calibration error did not differ between Jev and Opus 5.5, the registered comparator (difference 0.007; 95% CI -0.026 to 0.038); Opus 5 was better calibrated than both. On Semigran-45, no pairwise difference in accuracy or calibration was significant.

The direction of miscalibration differed between models (Figure 1). On DDXPlus, Jev was overconfident: in its highest-confidence bin (547 cases), mean confidence was 0.98 and accuracy 0.88. Opus 5.5 was underconfident: its accuracy exceeded its stated confidence in every bin above 0.5 (for example, 0.97 accuracy at 0.87 mean confidence). Similar calibration error therefore reflects opposite failures, which matter differently for triage: overconfidence suppresses review, underconfidence triggers unnecessary review.

**Table 3.** Accuracy and calibration by case set.

| Case set | Measure | Jev | Opus 5, reasoning off | Opus 5.5, reasoning on |
| --- | --- | --- | --- | --- |
| DDXPlus (n = 1,000) | Top-1 accuracy | 72.0% | 79.5% | 87.4% |
| DDXPlus | Top-3 accuracy | 88.3% | 96.4% | 97.7% |
| DDXPlus | Expected calibration error | 0.116 | 0.073 | 0.109 |
| DDXPlus | Brier score | 0.417 | 0.304 | 0.200 |
| DDXPlus | Divergence from reference differential | 0.544 | 0.495 | 0.461 |
| DDXPlus | True diagnosis given exactly 0% | 62 (6.2%) | 0 | 0 |
| Zandi (n = 80) | Top-1 accuracy | 88.8% | 91.2% | 88.8% |
| Zandi | Top-3 accuracy | 91.2% | 96.3% | 97.5% |
| Zandi | Expected calibration error | 0.065 | 0.122 | 0.146 |
| Zandi | Brier score | 0.203 | 0.129 | 0.156 |
| Zandi | True diagnosis given exactly 0% | 2 (2.5%) | 0 | 0 |
| Semigran (n = 45) | Top-1 accuracy | 84.4% | 88.9% | 93.3% |
| Semigran | Expected calibration error | 0.095 | 0.057 | 0.154 |

**Figure 1.**
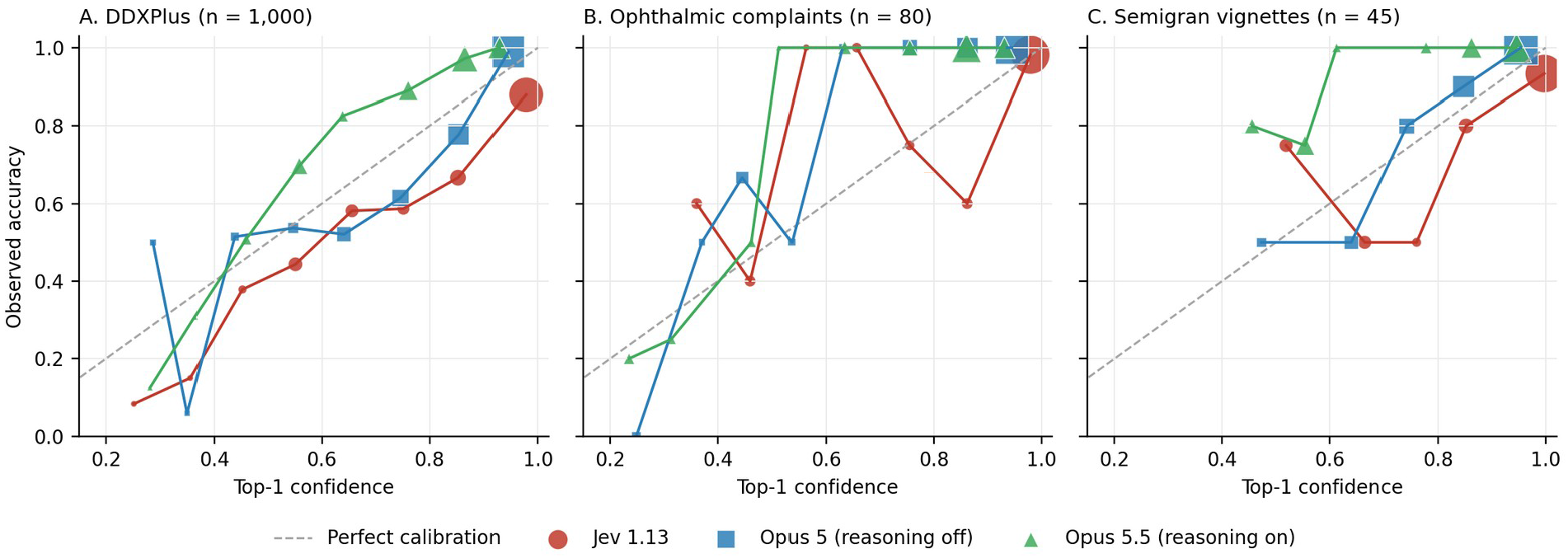
Calibration of each model’s top-1 answer. Points show mean confidence against observed accuracy in 10 equal-width confidence bins; marker area is proportional to the number of cases in the bin. Points below the dashed line indicate overconfidence; points above it indicate underconfidence.

Jev assigned exactly zero probability to the true diagnosis in 62 of 1,000 DDXPlus cases (95% CI 4.9% to 7.9%), and neither Claude model did so in any case (P<.001 for each paired comparison). Across all outputs, 92.0% of Jev’s probabilities were exactly zero and 98.8% fell on a 0.01 grid.

### Reasoning

Opus 5.5 reasoned before answering on 41.2% of DDXPlus cases (median 0 reasoning tokens; interquartile range 0 to 197) and on 10 of 45 Semigran cases. The accuracy gap between Opus 5 without reasoning and Opus 5.5 was 7.9 percentage points on DDXPlus.

### Uncertainty on uninformative inputs

On the 30 uninformative inputs with a forced choice, Jev placed a mean 58% on its top diagnosis, compared with 37% for Opus 5 and 21% for Opus 5.5. Its choices followed the option list rather than the input: vague eye complaints went to conjunctivitis (up to 93%), two vague vision complaints went to stroke (51% and 59%), and general complaints went to viral upper respiratory illness (up to 91%). When “none of these” was offered, Jev chose it for all 30 inputs (mean probability 0.82), compared with 77% of inputs for Opus 5 and 90% for Opus 5.5.

### Order sensitivity and absent diagnoses

Shuffling the option order changed all three models’ outputs slightly more than rerunning them did (mean excess TVD 0.025 for Jev, 0.034 for Opus 5, 0.019 for Opus 5.5; all P<.001), but rarely changed the top diagnosis. Jev did not differ from either comparator. When the true diagnosis was removed and “none of these” offered, all models chose it in 53% to 58% of cases; when they instead committed to a wrong diagnosis, Jev did so with the highest probability (mean 0.40 vs 0.29 to 0.34).

Jev’s and Opus 5’s top-1 errors on DDXPlus were moderately correlated (phi 0.57), more so than Jev’s and Opus 5.5’s (phi 0.46).

### Ophthalmic complaints and options-only baseline

On the 80 ophthalmic complaints, top-1 accuracy did not differ between Jev and either comparator (only Jev correct in 3 cases vs 5 for Opus 5, P=.73; 3 vs 3 for Opus 5.5, P>.99). Jev was better calibrated than Opus 5.5 (ECE difference -0.081; 95% CI -0.124 to -0.018) and did not differ from Opus 5 (-0.057; 95% CI -0.095 to 0.015). Its misses were harder misses: the true diagnosis fell outside its top 3 in 8.8% of cases (vs 3.8% and 2.5%), and it assigned exactly zero to the true diagnosis twice.

All three models were more accurate on the 5-descriptor version of each complaint than on the 3-descriptor version (Figure 3): Jev 82.5% vs 95.0% (P=.13), Opus 5 82.5% vs 100% (P=.02), and Opus 80.0% vs 97.5% (P=.02). On the sparse versions, the three models were equally accurate.

**Figure 2.**
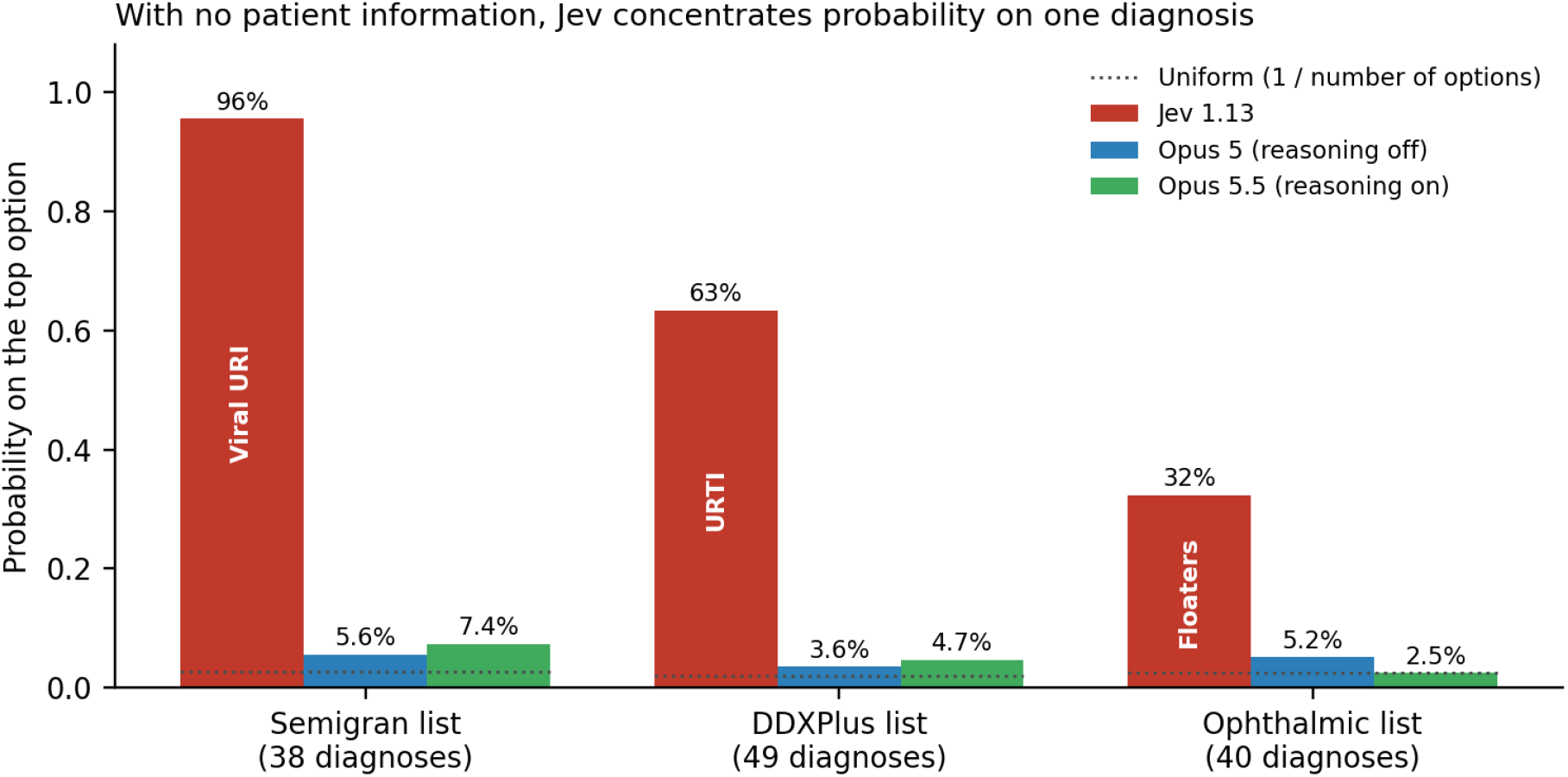
Probability each model placed on its top option when shown only the list of diagnoses, with no patient information. Dotted lines mark a uniform distribution over the list. Labels inside Jev’s bars name its choice.

**Figure 3.**
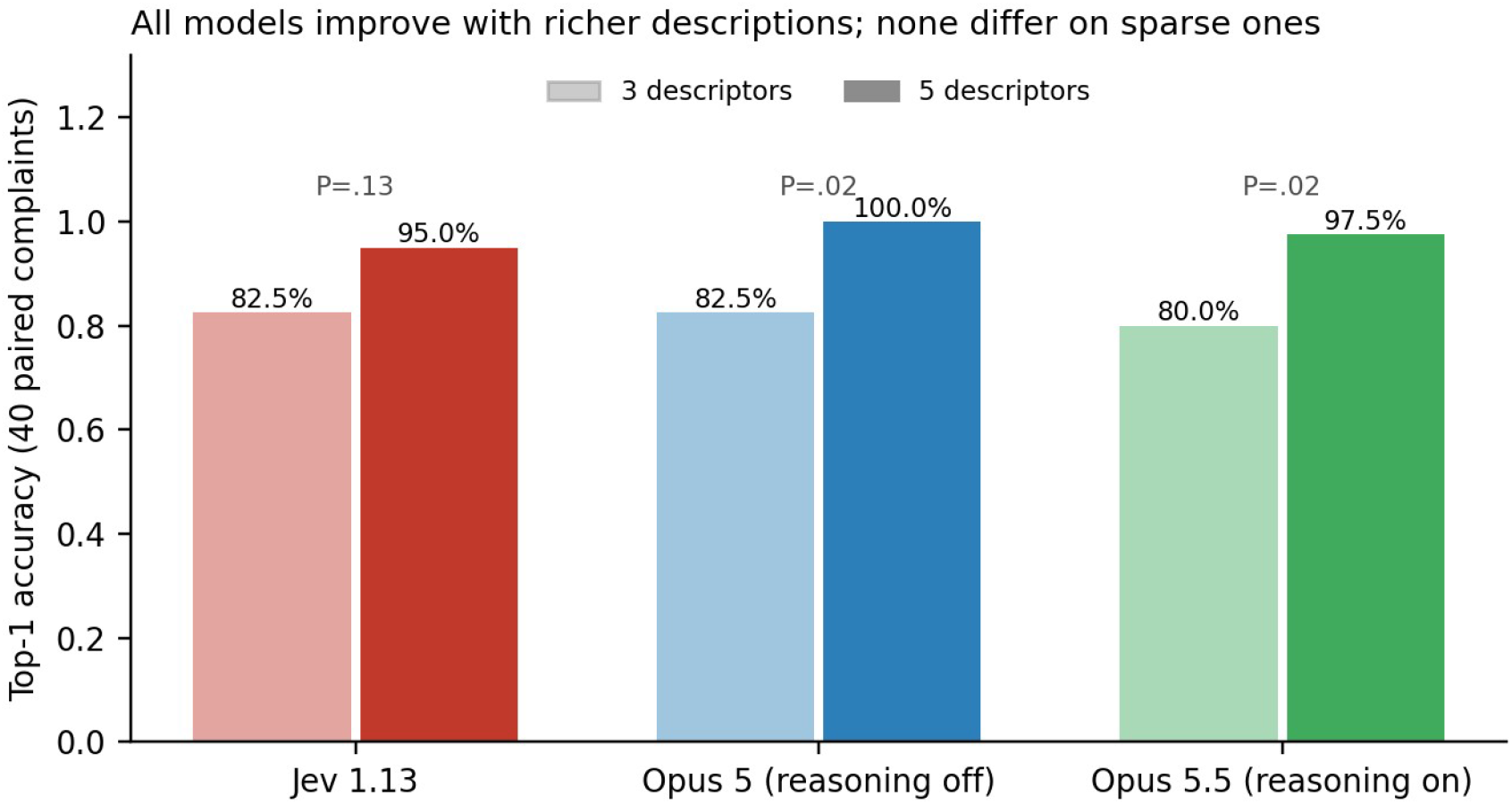
Top-1 accuracy on the 40 ophthalmic diagnoses written with 3 descriptors and with 5 descriptors. P values are exact McNemar tests within each model.

On the registered order test, Jev showed more violations than Opus 5 on this set (13 vs 4 cases; P=.049, not significant after correction for multiple primary comparisons). On the matched comparison, Jev was more order-sensitive than Opus 5.5 (excess TVD difference 0.015; 95% CI 0.005 to 0.025) and did not differ from Opus 5.

Given each option list with no patient information, Jev concentrated its probability on one diagnosis (Figure 2): viral upper respiratory illness at 95.6% for the Semigran list, upper respiratory tract infection at 63.4% for DDXPlus, and floaters at 32.4% for the ophthalmic list. The comparators spread probability almost evenly (normalized entropy 0.96 to 1.00, vs 0.07 to 0.66 for Jev). Applied to every case, no model’s options-only choice beat chance except where a diagnosis appeared twice on the Semigran list.

### Deviations from the registered plan

- The registered order-sensitivity criterion compared the largest of five permutation distances with a single-pair noise threshold, which flags about 23% of cases from noise alone. We report it as registered and add a matched comparison of permutation and repeat distances.
- Registered primary hypothesis H3 (irrelevant options) and secondary hypothesis H2 (paraphrases) were not run, because clinician-confirmed distractors and paraphrases were not prepared.
- The registered rule that Claude answer without reasoning could not be met for Opus 5.5, which does not allow reasoning to be disabled. We kept Opus 5.5 as the registered comparator and added Opus 5 with reasoning disabled.
- A sampled-answer arm for Opus 5.5 was dropped: its failures arose from reasoning exhausting a 50-token limit, and successful cases returned the same answer on all 20 samples.
- The Lyons ophthalmic set [16] was replaced by the registered Zandi set, because its vignette text is published only as an image. The options-only baseline and the “none of these” variant for uninformative inputs were added as exploratory analyses.
- A tie-breaking error that could register spurious top-1 changes under reordering was corrected before final analysis.
- An analysis error that counted the options-only calls as one extra case in paired comparisons was found and corrected before the results reported here; per-model accuracy and calibration were not affected.

## Discussion

### Principal findings

How the decision model compared depended on the kind of case. On short, distinctive ophthalmic complaints, Jev was as accurate as two frontier language models and better calibrated than the registered comparator. On 1,000 multi-symptom synthetic patients, it was clearly less accurate, trailing even Opus 5 with reasoning disabled by 7.5 points, so the gap there did not come from reasoning alone. Across all three sets, its calibration error was never worse than the registered comparator’s.

The most clinically relevant finding is the exact zero. In about 1 in 16 DDXPlus cases, Jev did not rank the correct diagnosis low; it ruled it out. A triage system that sends low-probability diagnoses to a clinician still catches a diagnosis ranked tenth. It cannot catch one assigned 0%, because nothing downstream will look for it. Neither comparator produced a single exact zero on the true diagnosis in 1,000 cases.

Put simply, a model that assigns exact zeros too readily is unsafe for triage. A diagnosis given even a small probability can still be flagged for review; a diagnosis given 0% is invisible to every downstream check.

The second finding concerns what the probabilities mean. Asked to choose among diagnoses for an input with no clinical content, Jev committed with a mean 58% confidence, following the structure of the option list. Offered “none of these,” it abstained on every such input, more decisively than either comparator. The model can recognize an uninformative input, but its probabilities carry that signal only when the question allows it. A high probability from a forced choice is therefore not evidence that the input supported the answer.

The options-only baseline sharpens this point. Shown only the list of diagnoses, Jev put 96% on viral upper respiratory illness, while both comparators spread their probability almost evenly. A probability that high, produced from no clinical information, is the clearest evidence that Jev’s forced-choice outputs mix what the case says with what the option list suggests. Yet the same model abstained on 97% to 100% of uninformative inputs when abstention was offered, more reliably than either comparator.

Miscalibration is a distinct hazard from inaccuracy [3]. Triage rules act on thresholds, such as escalating a case for same-day review when the probability of an urgent diagnosis exceeds a set level. An inaccurate but well calibrated model at least signals when it is unsure, so uncertain cases still reach a clinician. An overconfident model pushes wrong answers past those thresholds with the confidence that suppresses review. On DDXPlus, Jev was overconfident while Opus 5.5 was underconfident; at similar calibration error, Jev’s pattern is the more dangerous for triage, because underconfidence produces unnecessary referrals rather than missed ones.

This study does not establish why the decision model held up on the ophthalmic complaints, but several features of that set may contribute. Each complaint was short and usually contained one distinctive cue, such as a chemical splash or a curtain over the vision, which favors recognition over synthesis; on the sparse 3-descriptor versions, all three models were equally accurate. The DDXPlus cases instead require combining many findings, and our template rendering of them is stilted, which may disadvantage a single-pass model more than one that can reason. Finally, with 80 cases the ophthalmic comparison had wide confidence intervals, so parity there is compatible with a modest true difference in either direction.

### Relation to prior work

Early independent tests of Jev [7-9] reported nondeterminism, question-dependent miscalibration, and above-chance accuracy from answer options alone. Our results confirm run-to-run variation in a clinical setting (a third of repeated outputs differed) and show that option-list structure drives answers when the input is uninformative. Earlier vignette studies found high diagnostic accuracy for language models on Semigran-style cases, and one found their stated confidence reasonably calibrated by Brier score [5,6]; our Semigran results agree, and the set was too small and too easy to separate the models.

### Implications

For triage systems built on Jev, three practices follow from these results, and other decision models should be tested the same way before similar systems are built on them. First, always include an abstain option in the question, because the probabilities of a forced choice do not reveal uncertainty. Second, report the rate of exact zeros on known-correct answers, because a threshold-based safety net cannot catch them. Third, test calibration on the clinical task itself; a calibration claim from general workflow benchmarks did not transfer to diagnostic accuracy here.

A natural safeguard is to include a fixed list of can’t-miss diagnoses, such as giant cell arteritis or retinal detachment, in every query, so that the system explicitly weighs them even when the complaint seems minor. Our results argue against treating a 0% on such a list as reassurance, because 0% was exactly the output that excluded correct diagnoses here. A safer design asks about each can’t-miss diagnosis as a separate yes-or-no question with a deliberately low threshold for escalation.

The reasoning comparison also matters for cost. Opus 5.5 reasoned on only a minority of cases and gained 8 points of accuracy over Opus 5 without reasoning. Case-by-case reasoning may buy accuracy where it is needed without paying for it everywhere.

### Limitations

All cases were written, not spoken, and none came from real patients; DDXPlus patients are synthetic, and our rendering of their structured findings is stilted. The Claude models may have seen the public case sets in training, which could favor them; Jev’s training data are undisclosed. Semigran and the ophthalmic set were too small to detect moderate differences. Registered hypotheses on irrelevant options and paraphrases were not run. Opus 5.5’s resolved version carried no dated snapshot identifier. Jev’s 0.01 output resolution makes its noise and distance measures coarser than the comparators’.

Finally, all three models are commercial products that change; our results describe the versions tested.

Two further limitations bear on clinical use. The ophthalmic set comprised 80 simulated complaints, a small sample of uncertain applicability to real patient calls; evaluation on real triage messages with known outcomes is needed. And each case was presented as a single message, whereas real triage is a conversation in which the clinician asks follow-up questions. Jev cannot ask for clarification, and we did not test whether the language models’ ability to do so would change their accuracy.

## Conclusions

In this pre-registered evaluation, a decision model marketed as calibrated matched frontier language models on short ophthalmic complaints, with better calibration than the registered comparator, but was substantially less accurate on multi-symptom synthetic patients, where it excluded the correct diagnosis outright in about 6% of cases. Its probabilities expressed uncertainty only when the question allowed abstention, and it assigned high confidence to option lists with no case attached. Clinical deployment of decision models should require task-specific accuracy and calibration testing, explicit abstention options, and reporting of zero-probability rates on correct answers.

## Data Availability

All code, case lists, model outputs, and analysis results are available at https://github.com/avnish-deobhakta/jev-clinical-calibration. The pre-registered protocol is available at https://osf.io/fpmg3. Source datasets are publicly available from their original publications: DDXPlus (Fansi Tchango et al., NeurIPS 2022), the Semigran symptom-checker vignettes (Semigran et al., BMJ 2015), and the ophthalmic vignettes (Zandi et al., 2024). The full Semigran vignette text is not redistributed; the repository provides case identifiers and reference diagnoses only.

https://figshare.com/articles/dataset/DDXPlus_Dataset/20043374

https://doi.org/10.1136/bmj.h3480

https://doi.org/10.3390/bioengineering11020120

## Declarations

### Use of AI

The study harness, analysis code, and a first draft of this manuscript were written with assistance from Claude (Anthropic), a model family that also serves as the comparator in this study. To limit the influence of that relationship, the analysis plan and thresholds were pre-registered before data collection, every deviation is reported, all code and results are public, the first author verified all case transcriptions, and all authors reviewed the manuscript.

### Author contributions

AD: conceptualization, methodology, software, formal analysis, data curation, writing (original draft). JC, RWSC, SJ, AP, and TYTT: clinical interpretation, writing (review and editing). RBR: supervision, writing (review and editing). All authors approved the final manuscript.

### Ethics

No human participants or patient data were involved. All cases came from published or synthetic datasets, so institutional review and consent were not required.

### Data and code availability

The pre-registration is at osf.io/fpmg3. Code, per-call results, data manifests, and version history are available at https://github.com/avnish-deobhakta/jev-clinical-calibration. DDXPlus (CC BY 4.0) and Zandi et al. (CC BY 4.0) cases are redistributed with the code; Semigran vignettes are referenced by case identifier and must be obtained from the original publication.

